# The relationship between wild-type transthyretin amyloid load and ligamentum flavum thickness in lumbar stenosis patients

**DOI:** 10.1101/2022.03.23.22272831

**Authors:** Andy Y. Wang, Harleen Saini, Joseph N. Tingen, Vaishnavi Sharma, Alexandra Flores, Diang Liu, Michelle Olmos, Ellen D. McPhail, Mina G. Safain, James Kryzanski, Knarik Arkun, Ron I. Riesenburger

## Abstract

**Background:** One key contributor to lumbar stenosis is thickening of the ligamentum flavum (LF), a process still poorly understood. Wild-type transthyretin amyloid (ATTRwt) has been found in the LF of patients undergoing decompression surgery, suggesting that amyloid may play a role. However, it is unclear whether within patients harboring ATTRwt, the amount of amyloid is associated with LF thickness.

**Methods:** From an initial cohort of 324 consecutive lumbar stenosis patients whose LF specimens from decompression surgery were sent for analysis (2018-2019), 33 patients met the following criteria: (1) Congo red-positive amyloid in the LF; (2) ATTRwt by mass spectrometry-based proteomics; and (3) an available preoperative MRI. Histological specimens were digitized, and amyloid load quantified through Trainable Weka Segmentation (TWS) machine learning. LF thicknesses were manually measured on axial T2-weighted preoperative MRI scans at each lumbar level, L1-S1. The sum of thicknesses at every lumbar LF level (L1-S1) equals “lumbar LF burden.”

**Results:** Patients had a mean age of 72.7 years (range 59-87), were mostly male (61%) and white (82%); and predominantly had surgery at L4-L5 levels (73%). Amyloid load was positively correlated with LF thickness (R=0.345, p=0.0492) at the levels of surgical decompression. Furthermore, amyloid load was positively correlated with lumbar LF burden (R=0.383, p=0.0279).

**Conclusions:** Amyloid load is positively correlated with LF thickness and lumbar LF burden across all lumbar levels, in a dose-dependent manner. Further studies are needed to validate these findings, uncover the underlying pathophysiology, and pave the way towards using therapies that slow LF thickening.

## INTRODUCTION

Lumbar stenosis is one of the most common spine disorders afflicting older patients, and one key pathophysiological contributor is thickening of the ligamentum flavum (LF). Despite the ubiquity of lumbar stenosis, the underlying pathophysiological process is still poorly understood.^1^ There has been increasing interest in the role of amyloid, misfolded protein aggregates that commonly deposit in the ligamentum flavum of older individuals.^2, 3^ The amyloid deposited in LF is almost exclusively wild-type transthyretin amyloid (ATTRwt), a type derived from wild-type transthyretin precursor protein. ATTRwt is also found as a form of systemic amyloidosis. Many diagnosed ATTRwt patients present with heart failure due to cardiac amyloidosis, and many of these patients have an antecedent history of carpal tunnel syndrome.^4^ Recent research has suggested that patients with ATTRwt amyloidosis may also present with an antecedent history of lumbar stenosis, with amyloid deposition in the LF hypothesized as possibly leading to its thickening.^3, 5-8^ As ATTRwt is found in the LF of many older individuals who ultimately do not develop cardiac amyloidosis, it may be of interest outside of systemic amyloidosis as a factor in the pathophysiology of lumbar stenosis.

Our group has previously reported that lumbar stenosis patients harboring ATTRwt in their LF have thicker LF at the symptomatic level that required surgery, when compared to stenosis patients without ATTRwt.^9^ Subsequent analysis inclusive of asymptomatic lumbar levels also revealed an increased LF thickness in all lumbar levels when harboring ATTRwt.^10^ This total sum of lumbar LF thicknesses is termed lumbar LF burden. These studies demonstrate that the presence of ATTRwt in the LF is associated with an increase in LF thickness, when compared to lumbar stenosis patients who do not harbor ATTRwt. However, it is still unclear whether within patients harboring ATTRwt, whether the amount of amyloid is associated with a dose-dependent increase in thickness. Only one previous paper studied this, reporting a strong relationship between the transthyretin amyloid load and LF thickness.^8^ However, their method of quantifying amyloid deposits in histological specimens of ligamentum flavum acquired from lumbar decompression was not explained.

In this study, we aim to explore the relationship between transthyretin amyloid load and LF thickness in lumbar stenosis patients harboring ATTRwt through a precise machine learning quantification method. We have previously described this method as superior to existing methods that are subjective and non-reproducible.^11^ Through exploring the relationship between amyloid load and LF thickness, we hope to elucidate the role that ATTRwt deposition in LF has in LF thickening and lumbar stenosis.

## MATERIALS AND METHODS

### Patient selection

Between 2018–2019, we investigated 324 consecutive patients who underwent decompression surgery for lumbar stenosis. Inclusion in the study required meeting the following criteria: (1) LF specimen that contained Congo red-positive amyloid; (2) ATTRwt amyloid by mass spectrometry-based proteomics; and (3) an available preoperative MRI. Each LF specimen was stained with Congo red for apple-green birefringence under polarized light. The Congo red-positive amyloid deposits were typed by liquid chromatography tandem mass spectrometry (LC-MS/MS, Mayo Clinic Laboratories, Rochester, MN, USA) using previously published methods.^12^ By LC-MS/MS, all specimens were of ATTR type, and all lacked mutant peptides indicative of pathogenic mutations and thus were consistent with ATTRwt. Out of this entire cohort, 33 patients met the above criteria. Since some patients had multiple specimens stained, we include 46 LF histology slides in this paper. Congo red stained slides were scanned and digitized using VENTANA DP 200 slide scanner (Roche Diagnostics, Rotkreuz, Switzerland) at a 20x magnification and 1 focus layer.

### Amyloid Quantification

Amyloid deposits were quantified through the Trainable Weka Segmentation (TWS) plugin through Fiji/ImageJ as previously described.^11^ This machine learning segmentation method utilized small sets of human-directed annotations to learn and recognize characteristics of amyloid deposits. Classes of interest were selected to include amyloid, glass slide, calcifications, and tissue (Figure 1A). User-directed annotations on the digital specimens provided training data provided training data for TWS to build a classifier to recognize each of these classes, particularly the amyloid deposits (Figure 1B, 1C). The classifier was trained on 15 total specimens, each building upon the previous set of training data. Any additional annotation was added due to suboptimal amyloid recognition from application of the previous classifier. The final trained classifier was then applied all specimens without further annotations, including the initial specimens used to train the classifier. The TWS segmentation results were visually inspected for appropriate fit to the original images by three researchers. To calculate amyloid load in each sample, the amyloid pixel count was divided by the sum of the amyloid, calcifications, and tissue pixel counts (Figure 1E). When a patient had multiple specimens, the amyloid load of all available slides was quantified, and the arithmetic mean reported.

**Figure 1.**
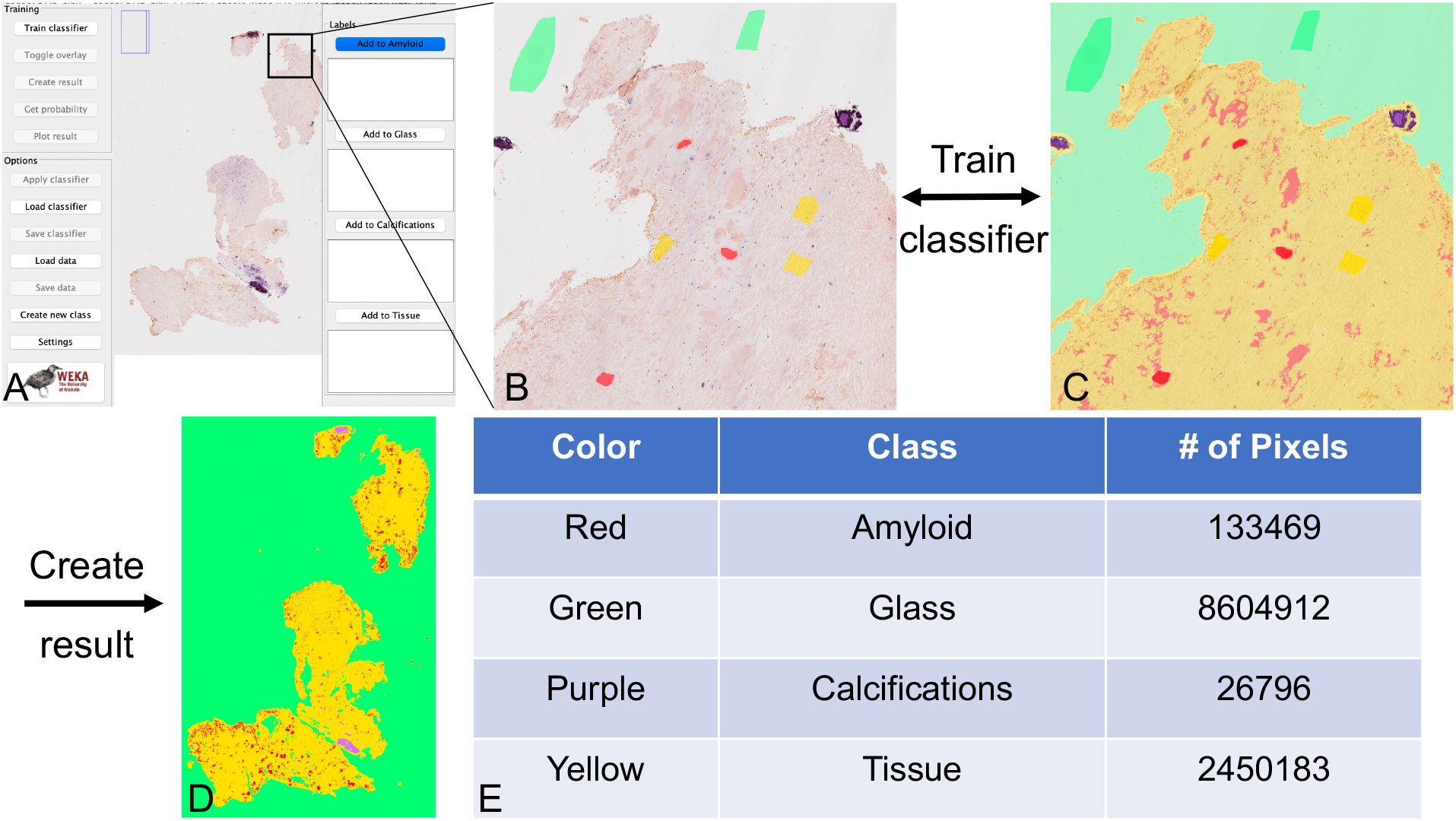
Quantification with TWS. A) A digitized image of the ligamentum flavum specimen is imported into Fiji and opened within the TWS graphical user interface. B) Manual annotations are drawn that correspond to each class of interest. C) The classifier is then trained to learn the characteristics of each class through these annotations and segments the rest of the image. An overlay is generated for the user to inspect the fit of the segmentation to the original image. Additional annotations may be added or prior annotations removed, re-training the algorithm as necessary until reaching desired fit. D) After satisfactory fit with the overlay, a final image is generated that assigns the pixels of each class to different colors. E) Calculation of the numbers of pixels in each class then allows for quantification of the total area of each component.

### Ligamentum flavum thickness measurements

Preoperative axial T2-weighted MRI scans were used to measure LF thickness at all lumbar levels (L1-S1). A sagittal view was used to localize the lumbar level involved, and LF thickness was measured in the MRI viewer at the facet joint of the lumbar level as previously described in Sakamaki et al.^13^ An example of how we quantified LF thickness is illustrated in Figure 2, showing perpendicular measurements at the edge of the inferior articular process of the facet joint. LF thickness was measured bilaterally at each level by measuring tools within the MRI viewer, and the arithmetic mean was recorded. Lumbar LF burden is calculated as the sum of mean LF thicknesses across all lumbar levels from L1-L2 to L5-S1. Two reviewers (who were uninvolved in the histological assessment of amyloid load) manually measured all specimens and were unaware of amyloid status. To assess inter-observer reliability, we generated an intraclass coefficient (ICC).^14^ Sample sizes for inter-rater reliability were determined using Walter et al. and Zou methods for estimations with ρ0 = 0.6 assumed for minimal acceptable value of reliability,^15, 16^ and calculations were carried out using a subset of 34 measurements. The ICC was found to be 0.89 (p < 0.0001, 95% CI 0.80–0.93), using ICC3. This finding is consistent with good inter-rater reliability (good: 0.75–0.9; excellent: > 0.9).

**Figure 2.**
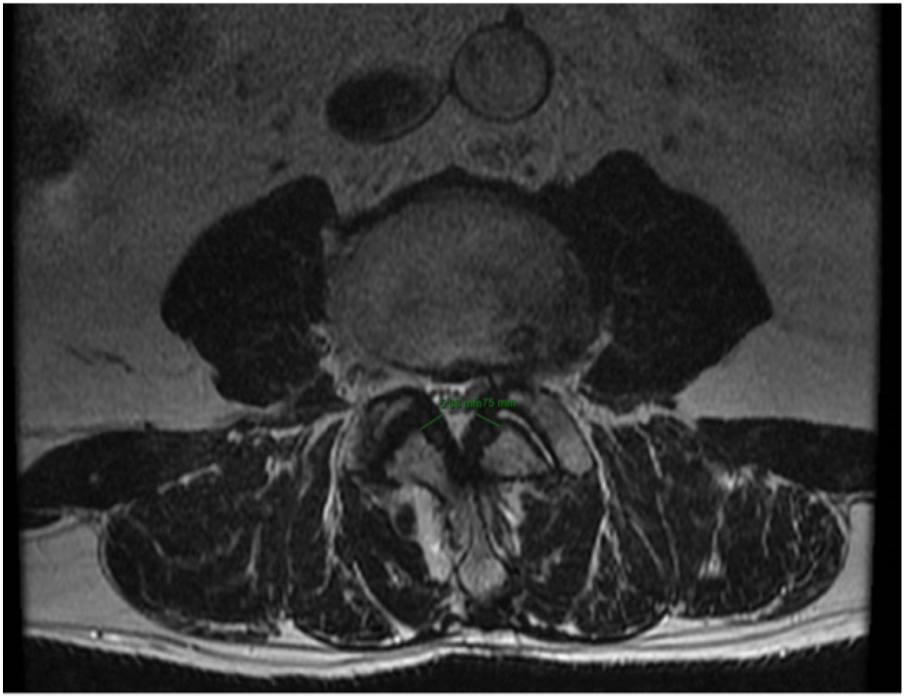
Example of LF thickness measurement on preoperative T2-weighted MRI.

### Statistical Analysis

Statistical analysis was conducted using R version 4.1.1 and figures were produced using the package *ggpubr*. Pearson correlation assessed the correlation between amyloid load and ligamentum flavum thickness, as well as between amyloid load and lumbar LF burden. A p-value < 0.05 was considered statistically significant.

## RESULTS

Baseline patient characteristics are listed in Table 1. The mean age of patients was 72.7 years old, with a standard deviation of 7.2 years (range 59-87). There was a preponderance of males (20 males vs 13 females). Most patients were white (82%). The level of decompression surgery, which could involve more than one level, showed a predominance at L4-L5 (73%).

**Table 1.** Baseline characteristics

| **Variable** | **Patients**<br>**N = 33<sup>1</sup>** |
| --- | --- |
| **Age** | 72.7 (7.2) |
| **Sex** |  |
| F | 13 (39%) |
| M | 20 (61%) |
| **BMI** | 29.7 (5.1) |
| **Level of Surgery** |  |
| L1-L2 | 4 (12%) |
| L2-L3 | 8 (24%) |
| L3-L4 | 13 (39%) |
| L4-L5 | 24 (73%) |
| L5-S1 | 5 (15%) |
| **Race** |  |
| Asian | 4 (12%) |
| African American | 1 (3.0%) |
| Hispanic | 1 (3.0%) |
| White | 27 (82%) |

<sup>1</sup>Mean (SD); n (%)

The percentage of amyloid load was calculated for each specimen, with a unimodal distribution and a peak between 3-4% (Figure 3A). LF thickness at the operative level was measured for each patient on preoperative MRIs, with the majority of thicknesses at similar levels between 3-7 mm (Figure 3B). The sum of all lumbar LF thicknesses, or “lumbar LF burden,” was calculated for each patient, and shown to have a bimodal distribution with peaks between 18-20 mm and between 26-28 mm (Figure 3C).

**Figure 3.**
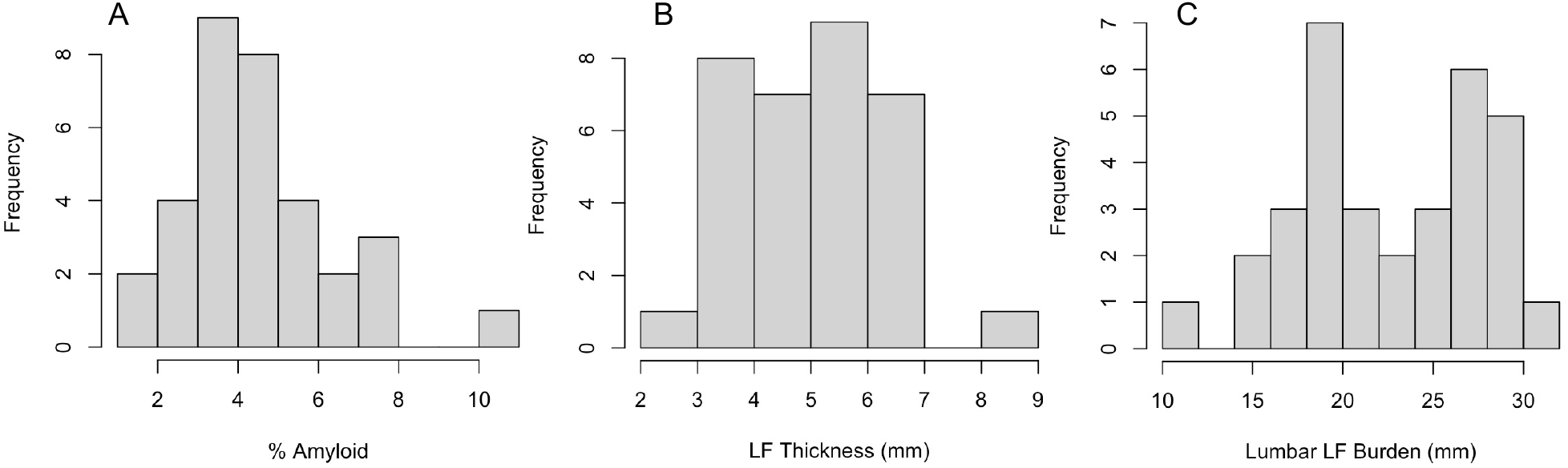
Histograms of A) amyloid load, B) LF thickness at the operative level, and C) lumbar LF burden.

A scatter plot of amyloid load and LF thickness at the surgical level is presented in Figure 4A. Amyloid load was shown to be positively correlated with LF thickness, achieving statistical significance (R = 0.345, p = 0.0492). Ligamentum flavum thickness was also measured between all lumbar levels L1-S1 including non-symptomatic and non-operative levels. A scatter plot of amyloid load and lumbar LF burden is presented in Figure 4B. Amyloid load was shown to be positively correlated with lumbar LF burden, achieving statistical significance (R = 0.383, p = 0.0279).

**Figure 4.**
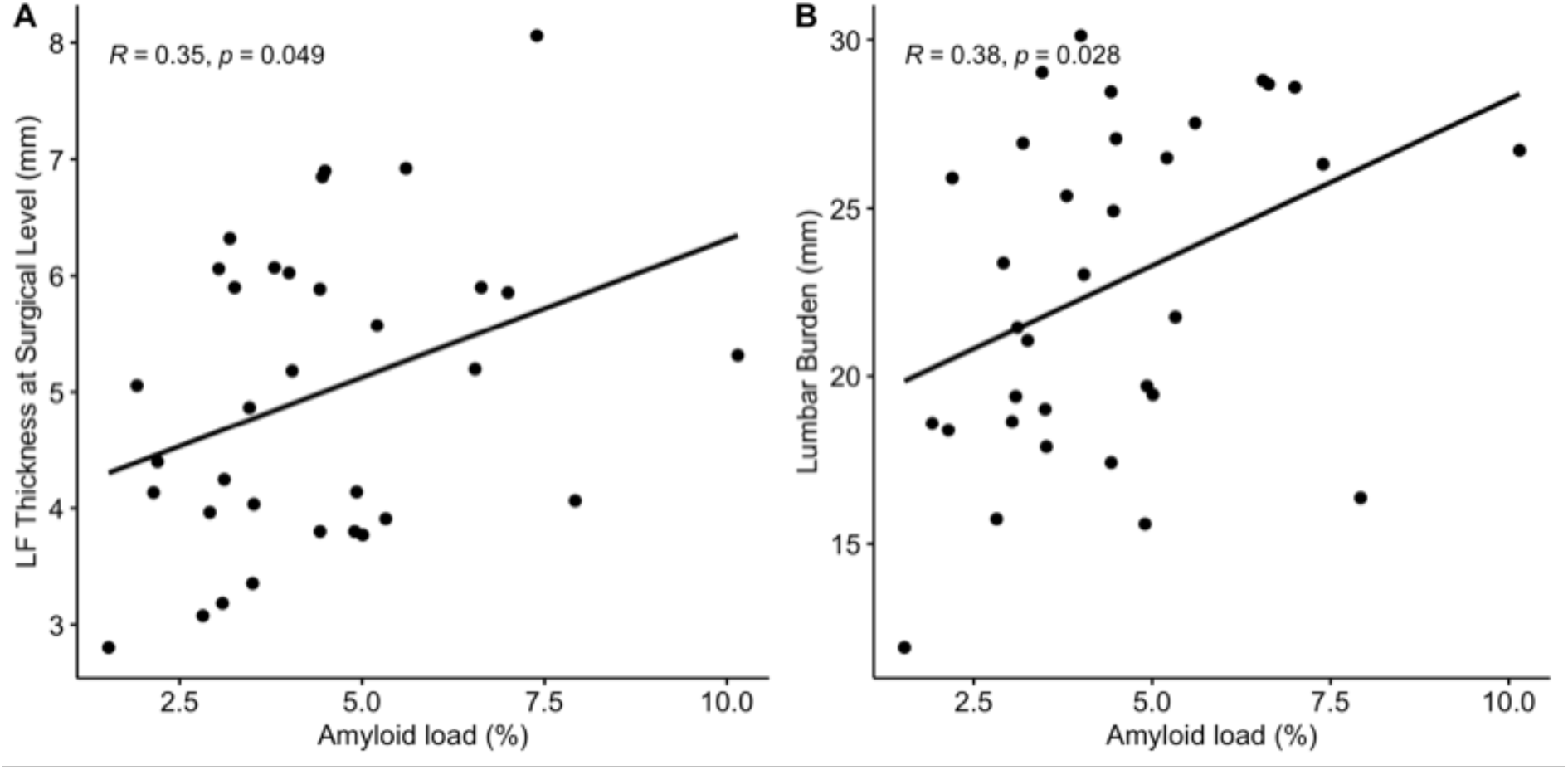
Scatter plots of amyloid load with surgical level LF thickness and lumbar LF burden. A) Amyloid load versus surgical level of LF thickness, positive correlation with p-value = 0.049. B). Amyloid load versus lumbar LF burden, positive correlation with p-value = 0.028.

## DISCUSSION

There is a positive and dose-dependent relationship between amyloid load and LF thickness at the surgical level as well as between amyloid load and lumbar LF burden. Lumbar LF burden is the sum of the LF thicknesses at all lumbar levels, and this positive association implies that the average LF thickness at each level is significantly correlated with amyloid load. These findings support a previous study by Yanagisawa et al. that analyzed 43 patients, finding positive correlation between amyloid load and lumbar LF thickness (r = 0.54, p-value < 0.0005).^8^ A limitation of that study was the methodology for obtaining amyloid deposit measurements, only reporting it was obtained with ImageJ without further detail. Other publications assessing amyloid load through computational methods have used the color thresholding function in ImageJ, which we have previously shown to be suboptimal.^11^ Our method utilizes powerful and precise machine learning quantification to measure amyloid load, which outperforms color thresholding. While the results shown in this paper are statistically significant, our p-values were not as low as those in the study by Yanagisawa et al. However, these p-values cannot be compared to each other as these are completely different studies. Furthermore, there may be a more complex relationship between the amount of amyloid and LF thickness in patients harboring ATTRwt in their LF and perhaps a larger sample size is needed for increased statistical power to better assess this relationship.

Our results suggest that amyloid may not simply be a bystander to LF thickening, but rather a direct contributor in a dose-dependent manner. The mechanism by which LF thickening occurs has yet to be fully understood. Currently, studies have identified TGF-β as part of the pathway in LF thickening, responsible for transforming fibroblasts to myofibroblasts and leading to increased collagen production.^17^ IL-6 is another implicated biomarker, associated with increased fibrosis and expression of collagen type I and III.^18^ TGF-β in the presence of IL-6 has been shown to induce an inflammatory response.^19^ Specifically, transforming growth factor beta (TGF-β) and interleukin 6 (IL-6) promote the differentiation of Th17 cells, leading to cytokine release. Additionally, the misfolding of amyloid itself could be another mechanism, with the normally buried hydrophobic residues now exposed to the environment.^20^ Future studies should focus on biochemical pathways of amyloid load within the ligamentum flavum in order to understand how amyloid specifically leads to LF thickening.

The study has several limitations. The presence of ATTRwt in one specimen was used to infer positivity at all levels, as the presence of amyloid cannot be confirmed at asymptomatic levels without further resection. There may also be sampling bias due to what the surgeon sends as a LF specimen. Some patients may have had previous spinal decompression surgeries at other levels prior to this study, underestimating the lumbar LF burden. Another limitation could be using a Congo red stain from a single block to extrapolate amyloid density throughout the specimen, as there may be variation in amyloid density in different blocks from the same specimen. Lastly, this study only includes patients undergoing decompressive surgery. Thus, there may be collider bias, also known as index event bias, which is when studies select patients based on an index event, in this case spinal surgery.^21^ This type of bias tends to shift study results towards the null, underestimating contributions as may be seen in this study.

In addition to understanding biochemical pathways, future studies may explore the potential of drug therapy for halting the progression of LF thickening in patients with amyloid. For ATTRwt, several therapies exist to reduce its production, stabilize ATTRwt, and remove fibrils, studied in the context of cardiac amyloidosis.^22^ Tafamidis is a well-known example that is FDA-approved for cardiomyopathy of wild-type or hereditary TTR-mediated amyloidosis.^23^ Although not yet studied in the context of LF thickening, tafamidis may potentially help reduce the buildup of ATTRwt in the LF and slow its thickening. This study lays the foundation for possible intervention studies such as a longitudinal study of patients on tafamidis, or even a clinical trial.

## CONCLUSIONS

The data from this study suggest that amyloid load is positively correlated with LF thickness in a dose-dependent manner in patients with ATTRwt. Further studies may investigate the biochemical pathways involved in this process, as well as potential drug targets for reducing LF thickening.

## Data Availability

Data in the present study are unavailable for release due to patient confidentiality.

## Acknowledgments

We would also like to thank Joanna R. O’Brien from the Tufts Pathology department for retrieving and scanning the histological slides of ligamentum flavum.

